# High feasibility of salivary therapeutic drug monitoring in linezolid, but less in tedizolid: A single-dose study in healthy subjects

**DOI:** 10.1101/2024.06.30.24309749

**Authors:** Hitoshi Kawasuji, Yasuhiro Tsuji, Keiko Miyaki, Takahiko Aoyama, Fumihiro Kurosaki, Masayoshi Ezaki, Yuki Koshiyama, Yusuke Takegoshi, Makito Kaneda, Yushi Murai, Kou Kimoto, Kentaro Nagaoka, Yoshihiro Yamamoto

**Author notes:** Corresponding author: Yoshihiro Yamamoto, MD, PhD Department of Clinical Infectious Diseases, Graduate School of Medicine and Pharmaceutical Sciences University of Toyama, 2630 Sugitani, Toyama 930-0194 Japan.

## Abstract

**Background:** Salivary therapeutic drug monitoring (TDM) offers the potential to reduce the risks, burden, time, and costs of blood-based TDM, but its feasibility in oxazolidinone antibiotics and the influence of food intake remain unknown.

**Methods:** A total of 12 healthy volunteers participated in this study. Linezolid and tedizolid were intravenously administered to 6 participants each. Saliva samples were taken at 15 time points and peripheral venous blood samples were also taken at 12 time points simultaneously with saliva. Total and unbound serum and saliva concentrations of linezolid and tedizolid were measured using high-performance liquid chromatography.

**Results:** Individual concentration–time curves in saliva versus serum (total and unbound) were similar in linezolid, but different in tedizolid. Saliva concentrations were significantly correlated with total and unbound serum concentrations in both agents. However, concentrations in each case and area under the concentration–time curve from 0 to 10 h (AUC_0–10_) in saliva were correlated with those in total or unbound serum for linezolid, but not for tedizolid. The mean saliva-to-serum (total and unbound) concentration and AUC_0–10_ ratios were 0.90 and 0.90 in total and 1.09 and 0.99 in unbound. Food intake did not influence these correlations in linezolid.

**Conclusions:** The analysis of linezolid in saliva is applicable for TDM as a promising alternative to conventional serum sampling without correlation factors, but application of tedizolid is less feasible. Easy sampling using a noninvasive technique may facilitate TDM even in underdeveloped countries with limited resources and specific patient categories.

## INTRODUCTION

Linezolid has been widely used in methicillin-resistant *Staphylococcus aureus* (MRSA) (1, 2) and vancomycin-resistant *Enterococcus faecium* (VRE) infections (3), and has recently gained a greater role in treatment regimens for multidrug-resistant or extensively drug-resistant *Mycobacterium tuberculosis*, or *Mycobacterium abscessus* (4–6). Patients should be closey monitored due to time- and concentration-dependent serious adverse effects of linezolid, including myelosuppression and neuropathy (7). In spite of linezolid being a drug with a very narrow therapeutic window (8), linezolid shows large pharmacokinetic variability, and drug–drug interactions also contribute to the high inter-individual variability in linezolid pharmacokinetics (9).

Therapeutic drug monitoring (TDM) serves as an efficient patient management tool by contributing to assessment of treatment response, helping to reduce toxicity and minimizing antibiotic resistance while ensuring adequate drug exposure (10). Several findings have provided evidence for the proper application of TDM for linezolid as an effective tool to predict serious adverse events and prevent discontinuation in advance (11–14).

Conventional venipuncture, the sampling currently used in clinical practice for TDM, is an invasive procedure with several logistical restrictions, such as the requirement of trained phlebotomists and appropriate materials, immediate storage in a refrigerator or freezer after collection, and cold chain transport to maintain the biospecimen integrity (15). Blood sampling is undesirable for some patient groups because of limited blood supply (e.g., neonates), less accessible veins (e.g., elderly), or religious objections (15). Alternatives to regular blood sampling (e.g., saliva) are therefore being studied.

Saliva sampling could reduce the risks, burden, time, and costs of blood sampling (16). Self-sampling at home would be advantageous, especially in TB-endemic countries, and would enable multiple sample collection (17). Dried blood spot (DBS) sampling is another less invasive method (18). However, DBS sampling can be painful, is more complicated, and has higher failure rates than saliva sampling (15). The drug concentrations in DBS are influenced by the hematocrit value, blood volume, sampling paper, and chromatographic effects (15, 18). In addition, unbound drug concentrations are not determinable in DBS; salivary concentrations are generally used to represent the unbound drug concentrations (15).

Serum contains unbound and bound drugs, whereas saliva generally contains only unbound drugs. Measuring the concentrations of unbound drugs may be important in pharmacokinetic studies, because only unbound drugs are pharmacologically active (19). This means that salivary concentrations of drugs may be more strongly associated with therapeutically active drug concentrations than total serum/plasma concentrations (19).

Previously, a few studies described linezolid concentrations in saliva and found that saliva is a suitable matrix for TDM in healthy individuals or MDR-TB patients (15, 17, 20–22). Salivary concentrations can be translated to plasma or serum concentrations with a correction factor of 1.0–1.2 (15, 20, 22). However, there has been no human study measuring unbound concentrations of linezolid and assessing the correlation between saliva and unbound serum/plasma concentrations. Only about 3–7 time points are generally used for linezolid measurement (15, 20, 21), and thus the data for linezolid pharmacokinetics in saliva and serum remain scarce.

Tedizolid is a second-generation oxazolidinone which has shown similar efficacy to linezolid in acute bacterial skin and soft tissue infections, with reduced adverse side effects (23, 24). Tedizolid has also shown high activity against *M. tuberculosis* including MDR strains (25) and species of nontuberculous mycobacteria (NTM) (26); thus, tedizolid is gaining recognition as an attractive alternative to linezolid for MDR-TB and NTM (27–29). Plasma protein binding differs between tedizolid and linezolid and is reported as 70–90% (29) and 18% (30) in humans; therefore the pharmacokinetics of these two drugs is considered to be different. However, there is scarce information regarding the pharmacokinetics in clinical practice. Furthermore, while there has been one study measuring salivary concentrations of tedizolid in rats (31), there has been no such study in humans.

In addition, salivary properties are known to influenced by food intake (16), but it is unclear whether diet may also affect salivary drug concentrations in linezolid or tedizolid. The aim of this study was to explore the feasibility of saliva-based TDM of linezolid and tedizolid in clinical settings, including the requirement for dietary control for saliva collection. To this end, we investigated the correlations of saliva and total and unbound serum concentrations in healthy volunteers, and assessed these correlations before and just after food intake.

### Participants and Methods

This study was registered at UMIN (UMIN000046556) and approved by the ethical review board of the University of Toyama (approval nos. R2012133 and R2020147) and the Nihon University (approval nos. 20-005 and 23A-005). Written informed consent was obtained from all participants.

A total of 12 healthy volunteers participated in this study. Linezolid was administered to 6 participants and tedizolid to 6 participants. Linezolid (Zyvox^®D^ IV bag 600 mg / 300 mL) and tedizolid phosphate (Sivectro^®D^ 200mg vial) for infusion were purchased from Pfizer (Tokyo) and MSD (Tokyo), respectively. Linezolid 600 mg/300 mL bag and tedizolid phosphate reconstituted with 4 mL of sterile water and further dissolved with 250 mL of 0.9% sterile saline were infused intravenously for 1 h.

Saliva samples were taken at 15 time points according to a preset schedule (**Table S1**) consisting of a sample before and at 0.25, 0.5, 0.75, 1, 1.25, 1.5, 2, 2.5, 3, 4, 5, 6, 8, and 10 h after the start of the infusion. To collect saliva samples, participants were asked to drink 50 mL of distilled water to promote saliva secretion before saliva collection. Subsequently, the saliva samples were collected by chewing on a cotton roll and processed using Salivette (Sarstedt, Nulmbrecht, Germany) in combination with centrifugation (3000×*g* for 2 min) according to the manufacturer’s instructions. Peripheral venous blood samples were also taken simultaneously with saliva, before and at 0.5, 1, 1.5, 2, 2.5, 3, 4, 5, 6, 8, and 10 h after the start of the infusion, through a short venous catheter inserted in the forearm opposite the infusion site. The serum samples were centrifuged at 3000×*g* for 10 min. All samples were stored at -80°C until analysis. During the study, participants ate the same food at the specified times (linezolid, food consumption between 4 and 5 h (4–5 h) after the start of the infusion; tedizolid, at 3–4, 6–8, and 8–10 h after the start of the infusion) and drank only water, which was unrestricted, from the start to the end of sampling.

Total and unbound serum concentrations of linezolid and tedizolid were measured using the improved methodology based on our previously published high-performance liquid chromatography (HPLC) methods (30, 32). Saliva samples were applied in the same manner as serum to measure saliva concentration.

Linezolid bulk powder (CAS No. 165800033) was purchased from Toronto Research Chemicals (North York, Canada), and tedizolid bulk powder (CAS No. 856867-55-5) was purchased from LKT Laboratories (Saint Paul, MN). Tedizolid was used as an internal standard (IS) for linezolid. L-tryptophan methyl ester hydrochloride was purchased from Tokyo Chemical Industry Co. (Tokyo) and used as an IS for tedizolid. All other reagents were of analytical grade and were commercially available. Linezolid and tedizolid concentrations were determined using an HPLC system (Shimadzu Corporation, Kyoto, Japan). High linearity was exhibited over a concentration range for linezolid and tedizolid (R^2^>0.999). The lower limit of quantifications (LLOQ) of linezolid and tedizolid were 0.1 and 0.001 mg/L, respectively. All concentration levels met the pre-set criteria for accuracy and precision (bias and coefficient of variation [CV] <15%; at LLOQ both <20%).

Area under the concentration–time curve from 0 to 10 h (AUC_0–10_) in serum and saliva was calculated using the trapezoidal rule. AUC_10-∞_ was determined using the equation C_10_/elimination rate constant (ke). ke was determined by using log-linear regression of the concentrations in the terminal period. AUC_10-∞_ was calculated by adding AUC_0–10_ and AUC_10-∞._ . Half-life (t_1/2_) was calculated using the equation 0.693/ke. C_max_ was defined as the highest observed concentration and T_max_ as the corresponding time of C_max_.

PK analyses, including determination of the C_max_, T_max_, AUC_0–10_, AUC_10-∞_, and t_1/2_ of linezolid and tedizolid, were performed in R (v4.3.3) applying the ggplot2 package (v3.5.0) (22) in a tidyverse framework (v2.0.0).

Passing–Bablok regression and Bland–Altman plots were used to analyse results. Pearson’s correlation and the Wilcoxon signed-rank test were applied to other comparisons. Statistical significance between different groups was defined as *P* <0.05. Statistical analysis and figure construction were performed using JMP Pro version 17.0.0 software (SAS Institute, Cary, NC) and GraphPad Prism version 9.5.1 (GraphPad Software, San Diego, CA). Data are expressed as mean with standard deviation (SD) or median with interquartile range (IQR).

## RESULTS

Participant demographics and the pharmacokinetic parameters (AUC_0–10_, AUC_10-∞_, C_max_, T_max_, t_1/2_, protein binding rate) of linezolid are shown in **Table 1**. Individual linezolid concentration–time curves in saliva versus serum (total and unbound) were similarly shaped (**Fig. 1A**) and T_max_ in saliva was not delayed (**Table 1**), which suggested that penetration of linezolid into saliva is fast. The linezolid AUC_0–10_, AUC_10-∞_, and t_1/2_ in saliva were similar to those in unbound serum, but C_max_ in saliva was similar to that in total serum rather than that in unbound serum.

**Figure 1.**
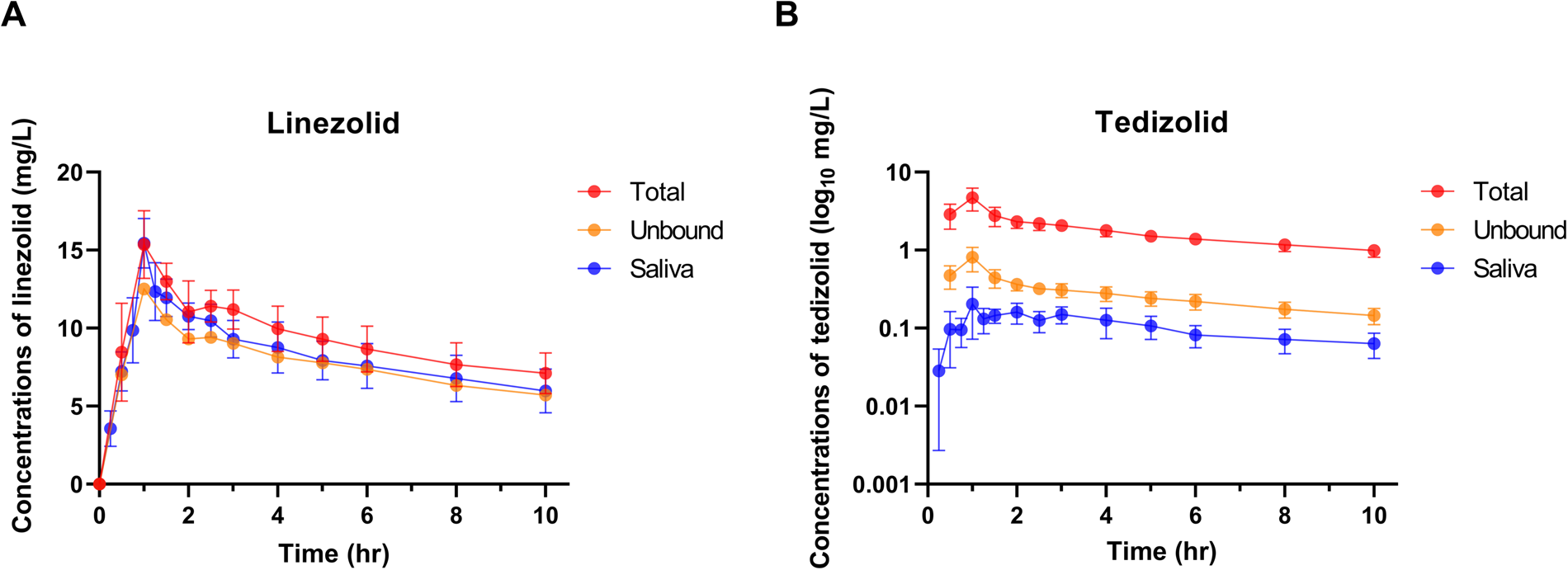
Concentration–time curves for linezolid and tedizolid in total and unbound serum and saliva **(A)** Concentration–time curves in total (red) and unbound serum (orange) and saliva (blue) for linezolid (n = 6). **(B)** Concentration–time curves in total (red) and unbound serum (orange) and saliva (blue) for tedizolid (n = 6). Data are presented as means and standard deviations.

**Table 1.**
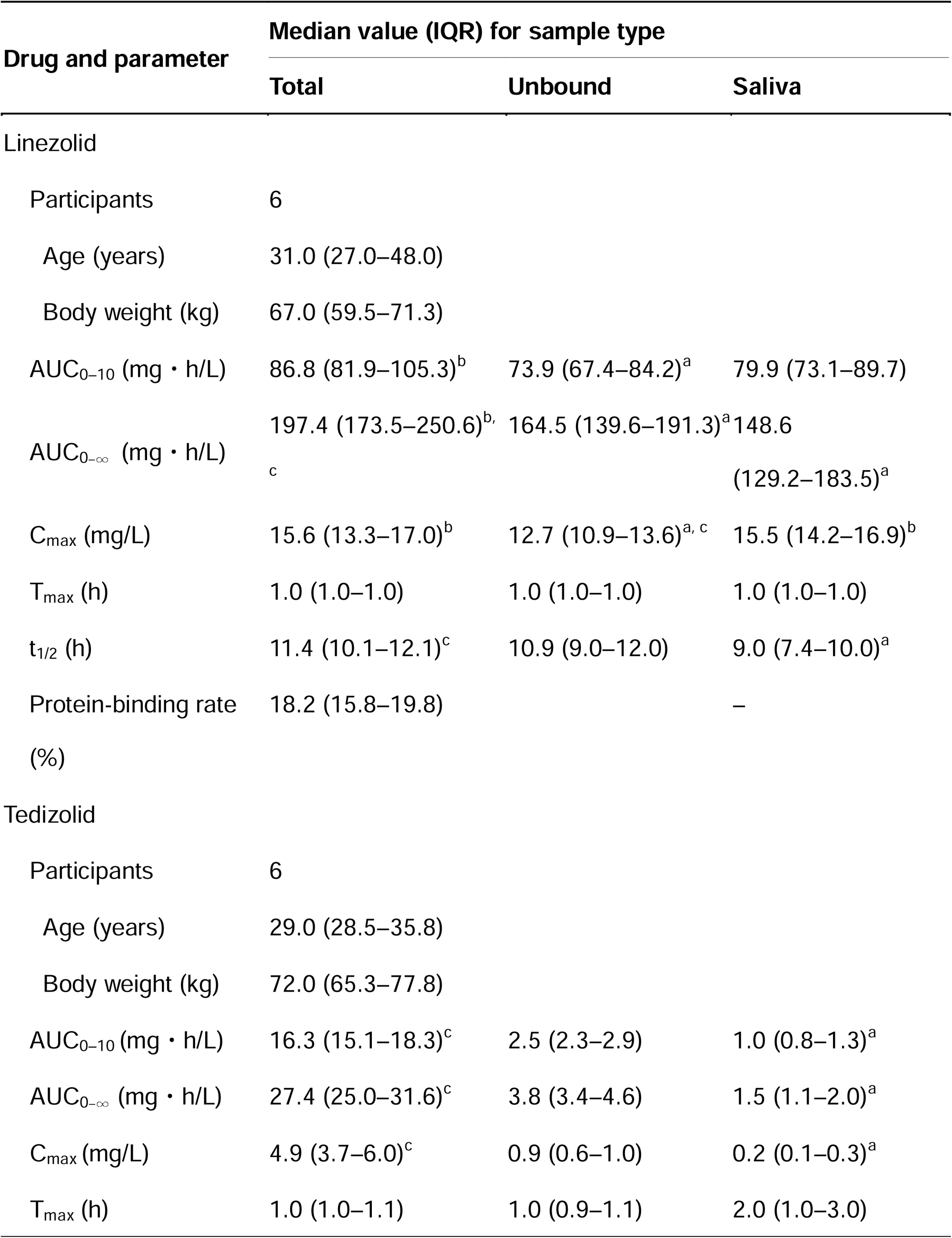
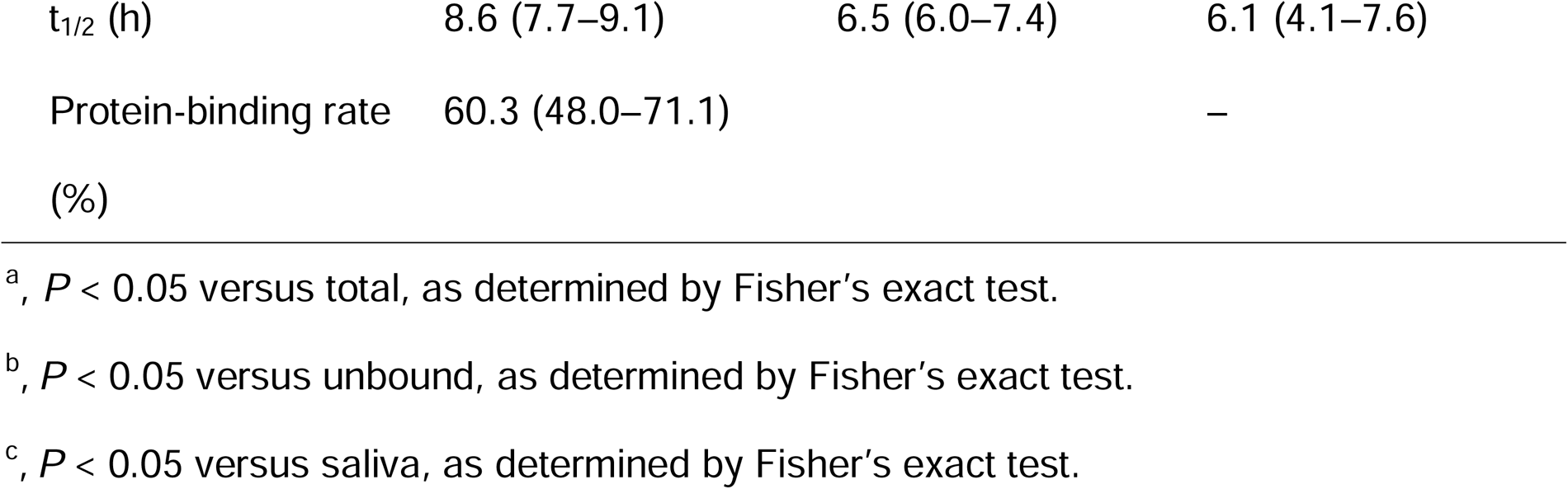
Pharmacokinetic parameters of linezolid and tedizolid in serum (total and unbound) and in saliva (n = 6)

Pearson’s test revealed that the total and unbound serum concentrations were significantly correlated with saliva concentration in total (**Fig. 2A and 2B**) and in each individual (**Fig. S1**) for linezolid. Passing–Bablok analysis also showed that the linear regression line of linezolid saliva concentration = -0.38 + 0.95 × total serum concentration with 95%CI of intercept -0.99–0.11; 95%CI of slope 0.88–1.02; R = 0.75, and *P* < 0.001. A linear relationship between unbound serum and saliva concentrations of linezolid was also observed: saliva concentration = -0.55 + 1.15 × unbound serum concentration with 95%CI of intercept -1.34–0.10; 95%CI of slope 1.04–1.25; R = 0.66, and *P* < 0.001, respectively.

**Figure 2.**
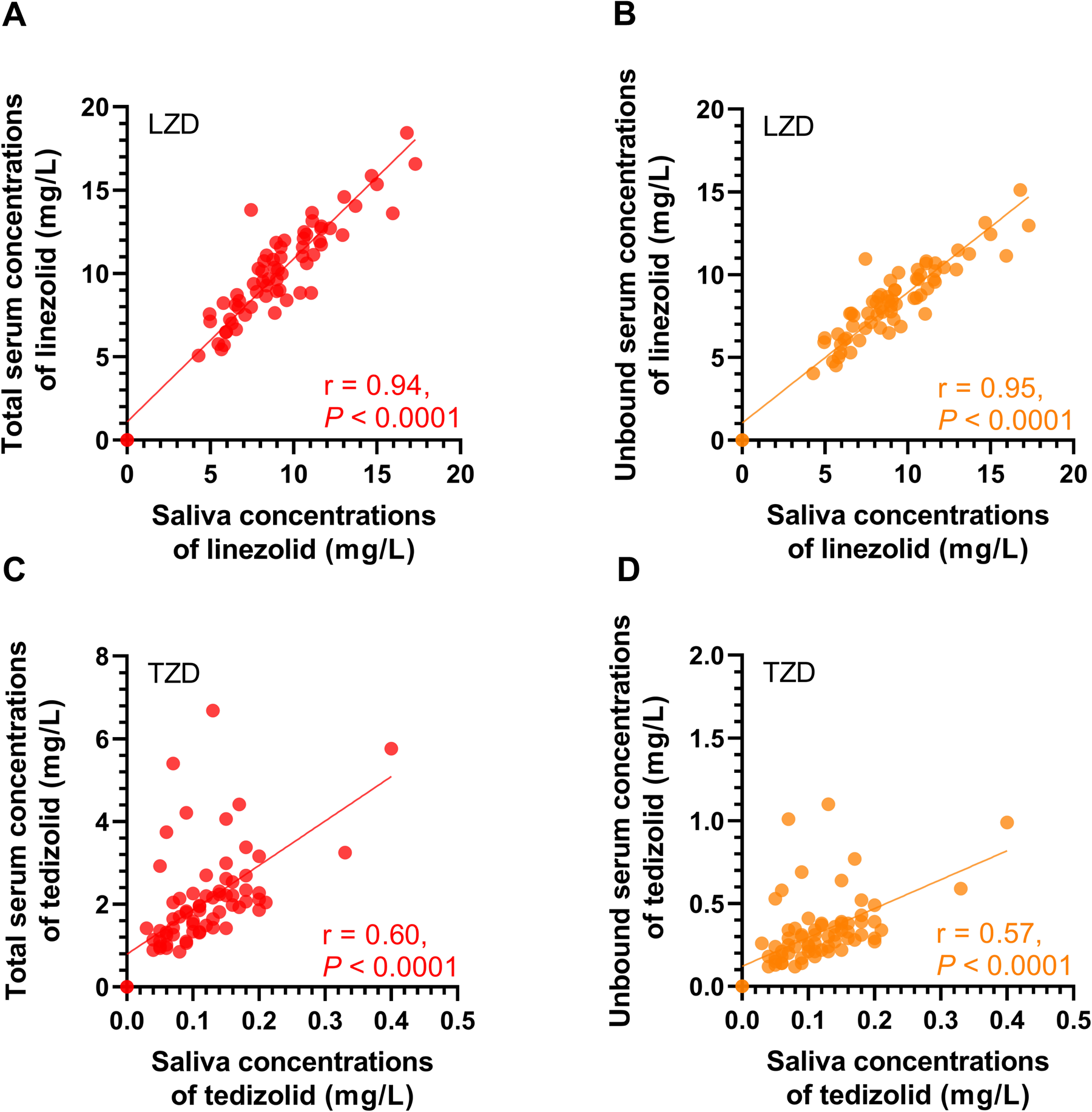
Correlation between total or unbound serum and saliva concentrations of linezolid and tedizolid The relationship between saliva concentrations and **(A)** total or **(B)** unbound serum concentrations of linezolid (n = 6). The relationship between saliva concentrations and **(C)** total or **(D)** unbound serum concentrations of tedizolid (n = 6). Each dot represents the concentration measured at one time point for one individual. The Pearson correlation was calculated, and the *P* value and r value and general linear regression lines are shown.

For AUCs, Pearson’s test also revealed that the total and unbound serum AUC_0–10_ and AUC_10-∞_ were correlated with saliva AUC_0–10_ and AUC_10-∞_, except for unbound serum AUC_10-∞_ (**Fig. 3A**). Passing–Bablok regression also showed a good relationship between the saliva AUC_0–10_ and total serum AUC_0–10_ with a slope estimate of 0.86 (95%CI, -0.33–2.86) and an intercept estimate of 5.44 (95%CI, -154.0–98.5), and unbound serum AUC_0–10_ with a slope estimate of 1.48 (95%CI, 0.14–2.52) and an intercept estimate of -26.6 (95%CI, -109.4–75.3), respectively.

**Figure 3.**
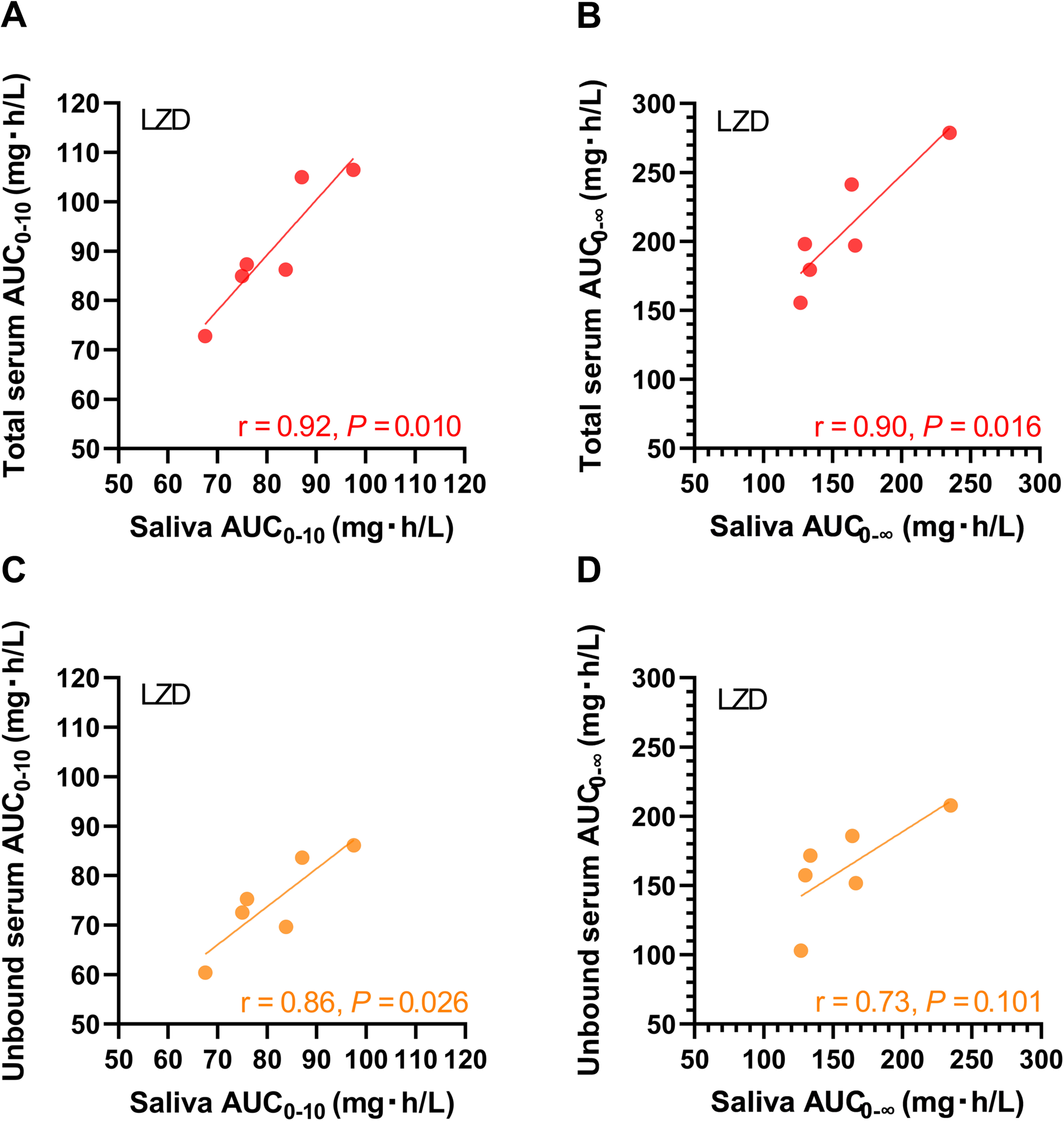
Correlation between saliva and total or unbound serum AUCs of linezolid. The relationship of area under the concentration–time curve from 0 to 10 h (AUC_0–10_) between in saliva and in **(A)** total or **(C)** unbound serum for linezolid (n = 6). The relationship of AUC extrapolated to infinity (AUC_10-∞_ ) between in saliva and in **(B)** total or **(D)** unbound serum for linezolid (n = 6). Each dot represents an individual value. The Pearson correlation was calculated, and the *P* value and r value and general linear regression lines are shown.

Bland–Altman assessment demonstrated good agreement between analyses of linezolid concentrations in saliva and serum (total and unbound). The mean saliva to total serum concentration, AUC_0–10_, and AUC_10-∞_ ratios were 0.90 (95%CI 0.67–1.13), 0.90 (95%CI 0.80–1.00), and 0.76 (95%CI 0.60–0.93), respectively (**Fig. 4A–C**), and the mean saliva to unbound serum concentration, AUC_0–10_, and AUC_10-∞_ ratios were 1.07 (95%CI 0.74–1.40), 1.09 (95%CI 0.94–1.23), and 0.99 (95%CI 0.63–1.35), respectively (**Fig. 4D–F**). Regarding the influence of diet, at 5 h after administration after food intake, the total and unbound serum concentrations were significantly correlated with saliva concentration (**Fig. S2**) and the biases (ranges) of saliva to total and unbound serum concentration ratios were 0.86 (95%CI 0.64–1.10) and 1.05 (95%CI 0.75–1.35), respectively. These results suggested that the correlation between serum and saliva may not be affected by food intake.

**Figure 4.**
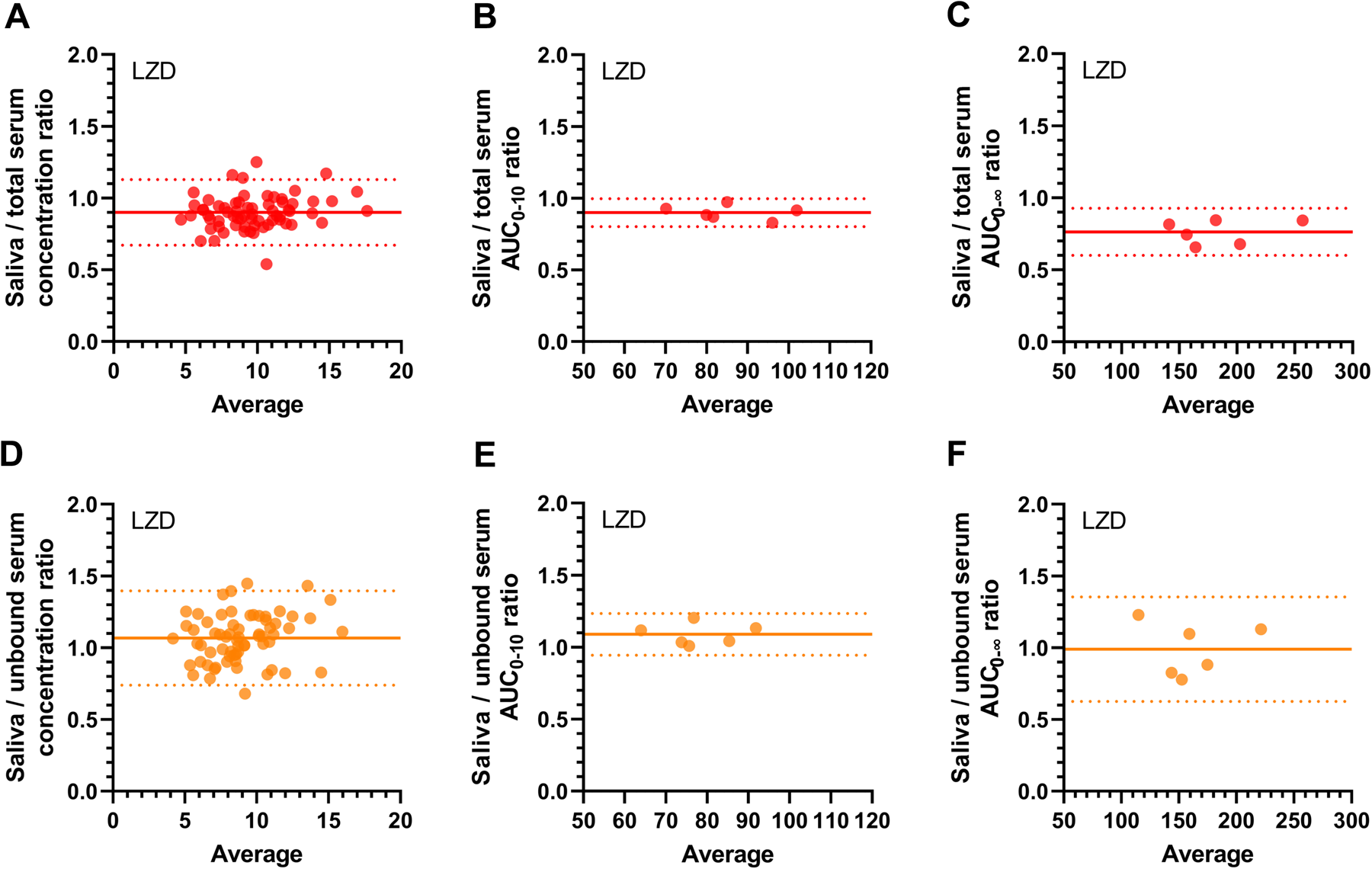
Bland-Altman plot of linezolid concentration, AUC_0–10_, and AUC_10-∞_ ratios in saliva versus total or unbound serum Bland-Altman plot of ratios of **(A)** concentrations, **(B)** AUC_0–10_, and **(C)** AUC_10-∞_ between saliva and total serum compared to the average of those in saliva and total serum. Bland-Altman plot of ratios of **(D)** concentrations, **(E)** AUC_0–10_, and **(F)** AUC_10-∞_ between in saliva and unbound serum compared to the average of those in saliva and unbound serum. Solid line: the bias. Dashed lines: the 95% limits of agreement.

Participant demographic and pharmacokinetic parameters of tedizolid are also shown in **Table 1**. Although the tedizolid AUC_0–10_, AUC_10-∞_, C_max_, and t_1/2_ in saliva were more similar to those in unbound serum than in total serum, individual tedizolid concentration–time curves in saliva versus serum (total and unbound) were differently shaped (**Fig. 1B**). Pearson’s test revealed that the total and unbound serum concentrations of tedizolid were significantly correlated with total saliva concentration (**Fig. 2C and 2D**). However, in the individual evaluations, 3 out of 6 participants showed no correlation between saliva and total or unbound tedizolid concentration (**Fig. S3 and S4**), which was due to the inconsistent results for T_max_ in saliva and serum; in 5 out of 6 participants (all but participant 2), peaks of saliva concentration were not observed and T_max_ in saliva was different from T_max_ in serum (**Fig. S3 and S4**). These difference seemed not to be due to the diet because T_max_ values in serum or saliva were measured before food intake (at 3–4, 6–8, and 8–10 h after the start of the infusion).

Passing–Bablok analysis showed that the linear regression line of tedizolid saliva concentration = 0.0055 + 0.053 × total serum concentration with 95%CI of intercept -0.0047–0.03208; 95%CI of slope 0.040–0.064; R = 0.54, and *P* < 0.001. The unbound serum concentrations of tedizolid were also linearly correlated with saliva concentrations with equation saliva concentration = 0.0067 + 0.36 × unbound serum concentration with 95%CI of intercept -0.005–0.021; 95%CI of slope 0.29–0.43; R = 0.50, and P < 0.001, respectively. However, for AUCs, Pearson’s test revealed that the total and unbound serum AUCs (AUC_0–10_ and AUC_10-∞_ ) were all uncorrelated with saliva AUCs (**Fig. S5**), which does not favour saliva as a sampling matrix for TDM. The mean (range) saliva to total and unbound serum tedizolid concentration rates were 0.060 (0.012–0.108) and 0.39 (0.07–0.74), respectively.

## DISCUSSION

This study is the first to investigate the relationship between saliva and unbound serum concentration with multiple time points and to measure saliva tedizolid concentrations in humans. We also investigated the influence of diet on the correlation between saliva and total or unbound serum concentrations, because while salivary properties are known to change according to food intake (16), the effect of diet on salivary drug concentrations has not been unclarified.

Saliva contains only the unbound fraction of drugs, since only the unbound fraction is able to infiltrate through the salivary tissues, including the capillary wall, the basement membrane, and the membrane of the salivary gland epithelial cells (33). Thus, salivary concentrations generally represent the unbound drug concentrations (15). Regarding linezolid, as we expected, the time-courses of linezolid saliva concentrations were similarly shaped with unbound concentrations in serum, and the mean saliva-to-serum concentration, AUC_0–10_, and AUC_10-∞_ ratios were closer to 1 for those to unbound serum compared to those to total serum. The mean saliva to unbound serum concentration, AUC_0–10_, and AUC_10-∞_ ratios were 1.07, 1.09, and 0.99, respectively (**Fig. 4D–F**).

To date, no human study has compared linezolid saliva and unbound concentrations in serum or plasma. Based on the previous studies conducted in TB patients, Bolhuis et al. reported the mean oral fluid/total serum concentration or AUC from 0 to 12 h (AUC_0-12_) ratios were 1.03 and 0.97, respectively (20). Van den Elsen et al. also showed that the median saliva/total serum concentration and AUC_0-24_ ratios were 0.76 and 0.81, respectively (17). They also suggested a correlation factor of 1.2 when translated to serum AUC_0-24_ using saliva AUC_0-24_, which may be due to the difference between the total and saliva concentrations, the latter of which represent the unbound concentrations, and it seemed that the values of saliva AUC_0-24_ and unbound serum AUC_0-24_ would be approximately the same. In the present study, the mean saliva-to-total serum concentration, AUC_0–10_, and AUC_10-∞_ ratios were 0.90, 0.90, and 0.76, respectively (**Fig. 4A–C**), and the mean saliva-to-serum concentration, AUC_0–10_, and AUC_10-∞_ ratios were closer to 1 for unbound serum concentrations than for the total serum concentrations. Although there were slight differences in saliva-to-total serum concentrations or AUC ratios in these studies, which might be attributable to differences in sampling methods, processing or storage, salivary TDM of linezolid indeed might indeed be feasible with good reproducibility and is ready for validation in a clinical setting.

Salivary properties are known to change according to food intake (16). To improve the feasibility of salivary linezolid TDM, we investigated the effects of food intake on salivary linezolid concentrations and the correlation with total and unbound serum concentrations. In the present study, even after food intake, total and unbound serum concentrations were significantly correlated with saliva concentration (**Fig. S2**) and the biases (ranges) of saliva to total and unbound serum concentration ratios were 0.86 (0.64–1.10) and 1.05 (0.75–1.35), respectively. These results suggested that the correlation between serum and saliva linezolid concentrations may not be affected by diet, and dietary control may not be necessary when collecting saliva.

For tedizolid, the mean (range) tedizolid saliva-to-unbound serum concentration ratio was 0.39 (0.07–0.74), suggesting lower passage into the saliva than linezolid. As in previous studies (29, 32), the serum protein-binding rates were different between linezolid and tedizolid in the present study. The median (IQR) serum protein-binding rates are 18.2 (15.8–19.8) in linezolid and 60.3 (48.0–71.1) in tedizolid, respectively (**Table 1**). It is thought that the significant difference in serum-protein binding between linezolid and tedizolid affects drug passage into the saliva. In addition to serum-protein binding, pH and pKa, lipid solubility, charge, molecular weight and spatial configuration, dose and clearance of the drug, salivary flow rate and pH, and salivary-binding proteins and salivary enzymes capable of metabolizing the drug would also affect the results (16, 34). Based on these many variables, the analysis of oxazolidinone concentrations in saliva and serum must be validated in humans, not animals. In fact, however, there has been only one study measuring salivary concentrations of tedizolid, and that study used a rat model; the results showed that the serum-protein binding and salivary pH values were different from those of humans (31). The study also demonstrated a strong correlation between saliva and plasma concentrations of tedizolid and suggested that saliva is a useful matrix for TDM of not only linezolid but also tedizolid (31). However, in the present study in humans, although correlations between saliva and total and unbound serum tedizolid concentrations were observed in participants overall, these correlations were not observed in 3 of the 6 participants (**Fig. S3**). A possible explanation for the observed nonlinear relationship might be the low tedizolid concentrations, higher interindividual variability, and the absence or delayed saliva concentration peak after infusion (**Fig. S3 and S4**). In addition, the total and unbound serum AUCs (AUC_0–10_ and AUC_10-∞_) were all uncorrelated with saliva AUCs in tedizolid (**Fig. S5**). Based on these results, saliva seems not to be useful in TDM for tedizolid.

Salivary TDM could be an attractive alternative method for traditional linezolid TDM using plasma or serum, and feasibility improvements will likely be a focus of future studies, including stability studies for transport at room temperature and cross-validation of existing analytical methods in saliva (17). However, with respect to tedizolid our data do not support saliva as a suitable matrix for TDM using the described method. As shown in a previous systematic review (15), saliva will likely not be a universal but only a selective matrix for TDM.

Saliva sampling is easy and noninvasive, and requires only a small sample volume (100 μL) for measurement, thus allows for the collection of multiple samples, while avoiding the risks associated with blood drawing. As with other drugs reported in previous studies (15, 35), the linezolid concentrations in saliva represent the unbound concentrations in the serum, and therefore salivary concentrations may be considered more closely related to therapeutically active unbound concentrations at the site of action than total serum concentrations. Saliva sampling might even reduce costs due to the higher level of training of personnel needed for blood sampling and because less time is needed. Moreover, saliva sampling might even take place at home. If collected saliva is stable for a certain period (e.g., a few weeks) even under low or high temperature, salivary TDM would allow children, elderly, and people with disabilities the option to sample themselves at any location and afterward bring their saliva samples to a local health post (15). To further improve feasibility of salivary TDM in clinical settings, the applicability of saliva and/or collection devices other than the Salivette (Sarstedt, Leicester, United Kingdom) for pharmacokinetic analysis and therapeutic drug monitoring in patients should be clinically validated (20).

There were several limitations in the present study. First, salivary pH values which could influence drug penetration into saliva were not measured. Second, we included only healthy volunteers in the present study and did not investigate children or patients, especially those with disease affecting the saliva composition. Third, we did not evaluate the time-stability of saliva samples under room temperature conditions.

In conclusion, the analysis of linezolid (with no correction factor) in saliva is applicable for TDM as a promising alternative to conventional serum sampling. Easy sampling using a noninvasive technique may facilitate therapeutic drug monitoring for specific patient categories.

## Transparency declaration

### Data availability

The data presented in this study are available upon request from the corresponding author.

### Author contributions

Conceptualization: H.K., Y. Tsuji, and Y.Y.; Methodology: H.K. and Y. Tsuji; Validation: H.K. and Y. Tsuji; Formal Analysis: H.K. and Y. Tsuji; Investigation: H.K., Y. Tsuji, K.M., T.A., F.K., M.E., Y.K., Y. Takegoshi, M.K., Y.M., K.K., and K.N.; Data Curation: H.K. and Y. Tsuji; Writing–Original Draft Preparation: H.K. and Y. Tsuji; Writing–Review and Editing: H.K. and Y. Tsuji; Visualization: H.K. and Y. Tsuji; Supervision: Y. Tsuji and Y.Y.; Project Administration: Y. Tsuji and Y.Y.; Funding acquisition: H.K. and Y.Y.

## Acknowledgments

We thank Kai Kurihara, Risa Sakurai, and Rei Yasukochi for their assistance with the sample collections.

---

**Table S1.**
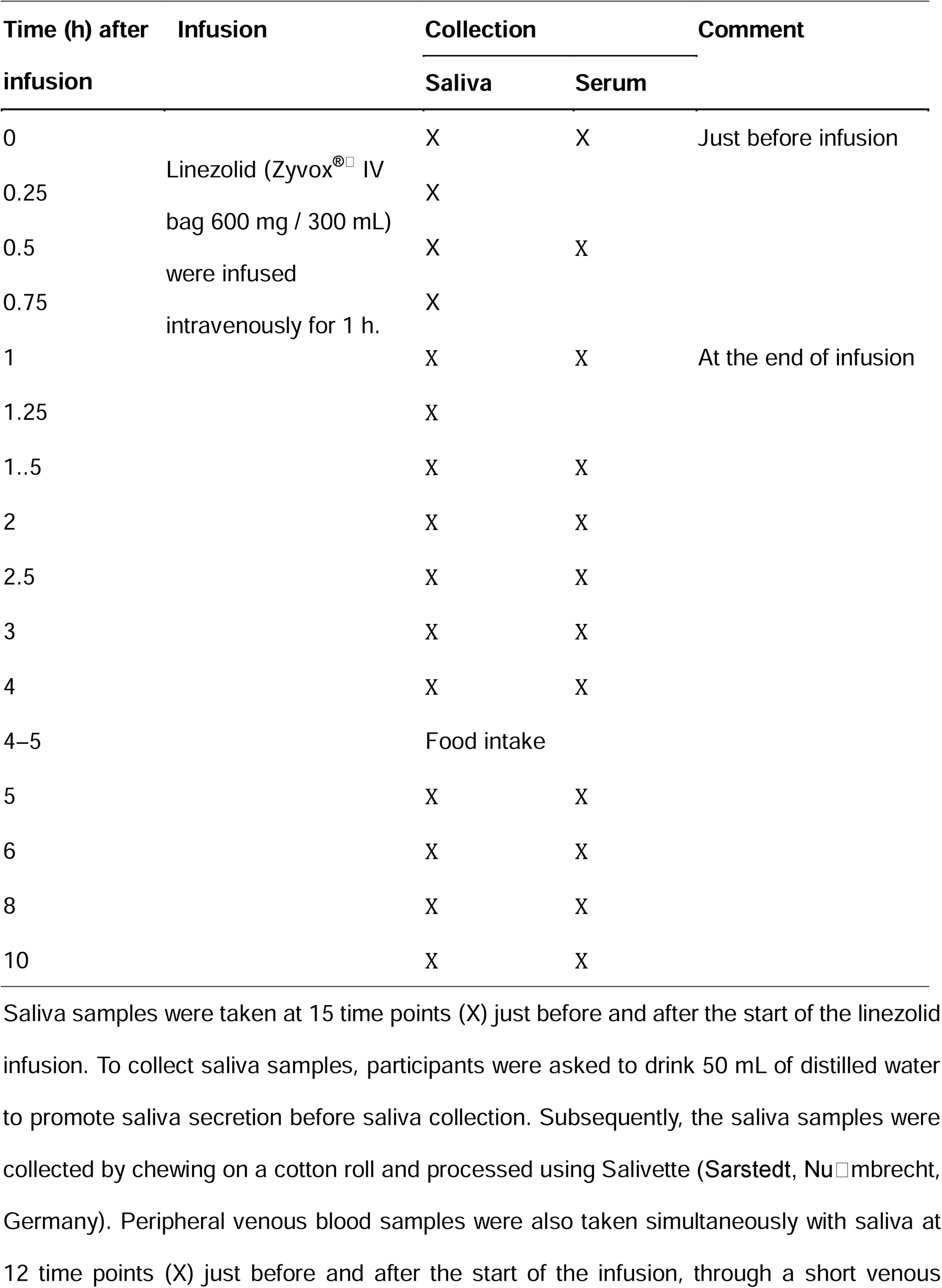
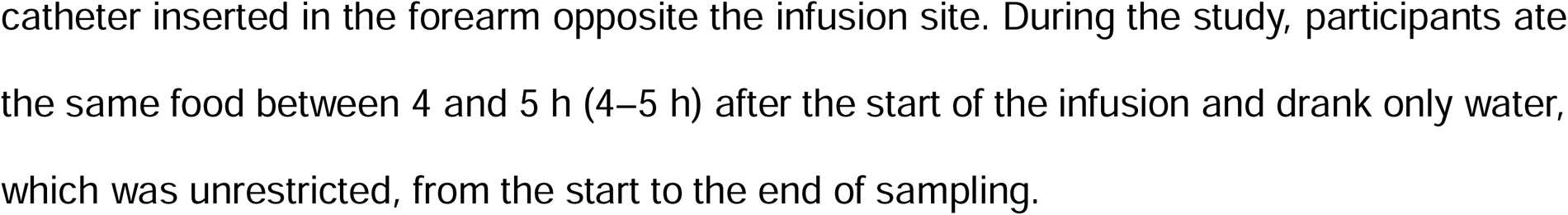
Time schedule of linezolid study.

| Time (h) after<br>infusion | Infusion | Collection |  | Comment |
| --- | --- | --- | --- | --- |
|  |  | Saliva | Serum |  |
| 0 | Linezolid (Zyvox® IV<br>bag 600 mg / 300 mL)<br>were infused<br>intravenously for 1 h. | X | X | Just before infusion |
| 0.25 |  | X |  |  |
| 0.5 |  | X | X |  |
| 0.75 |  | X |  |  |
| 1 |  | X | X | At the end of infusion |
| 1.25 |  | X |  |  |
| 1.5 |  | X | X |  |
| 2 |  | X | X |  |
| 2.5 |  | X | X |  |
| 3 |  | X | X |  |
| 4 |  | X | X |  |
| 4–5 |  | Food intake |  |  |
| 5 |  | X | X |  |
| 6 |  | X | X |  |
| 8 |  | X | X |  |
| 10 |  | X | X |  |

Saliva samples were taken at 15 time points (X) just before and after the start of the linezolid infusion. To collect saliva samples, participants were asked to drink 50 mL of distilled water to promote saliva secretion before saliva collection. Subsequently, the saliva samples were collected by chewing on a cotton roll and processed using Salivette (Sarstedt, Nußmberg, Germany). Peripheral venous blood samples were also taken simultaneously with saliva at 12 time points (X) just before and after the start of the infusion, through a short venous
catheter inserted in the forearm opposite the infusion site. During the study, participants ate the same food between 4 and 5 h (4–5 h) after the start of the infusion and drank only water, which was unrestricted, from the start to the end of sampling.

**Table S2.**
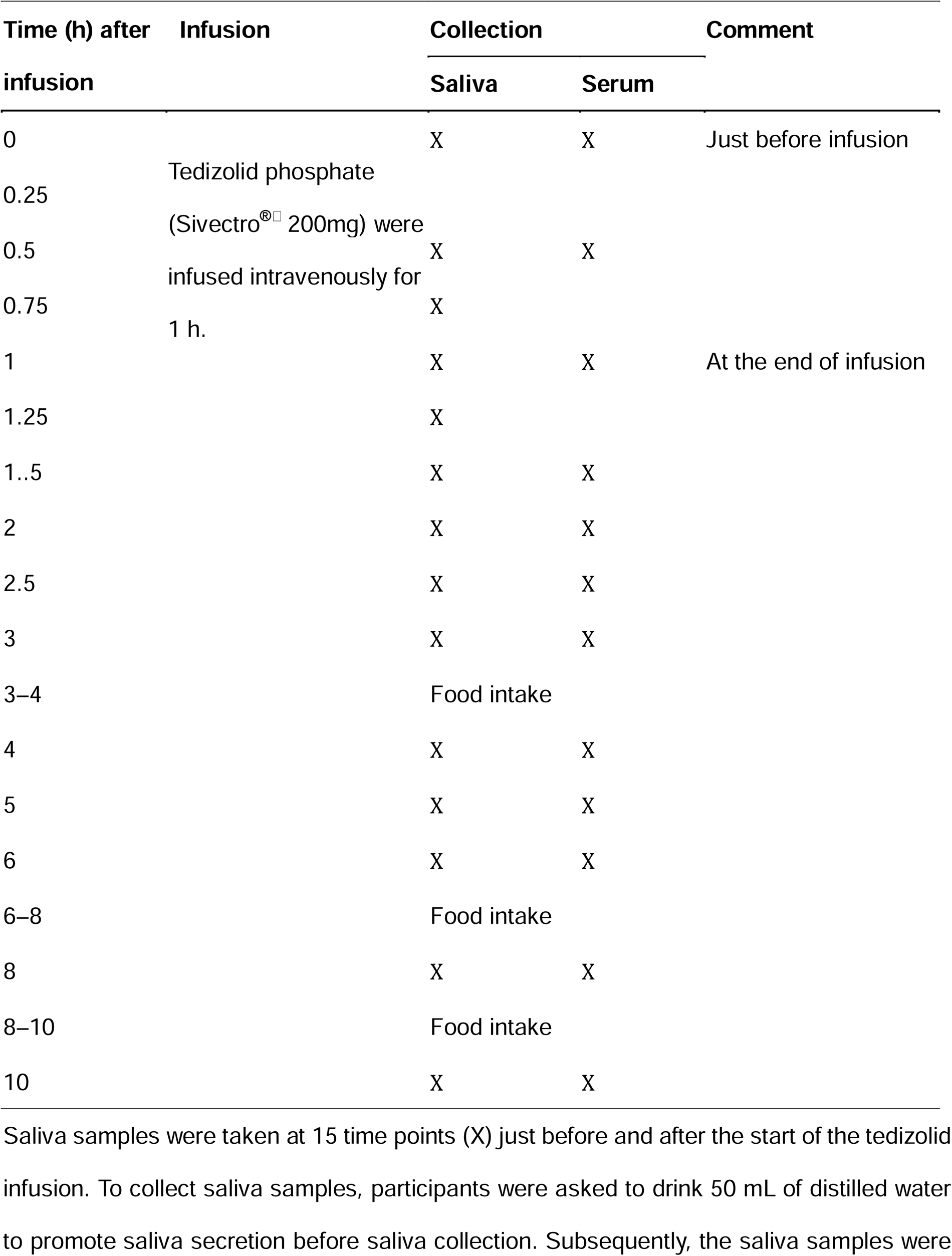
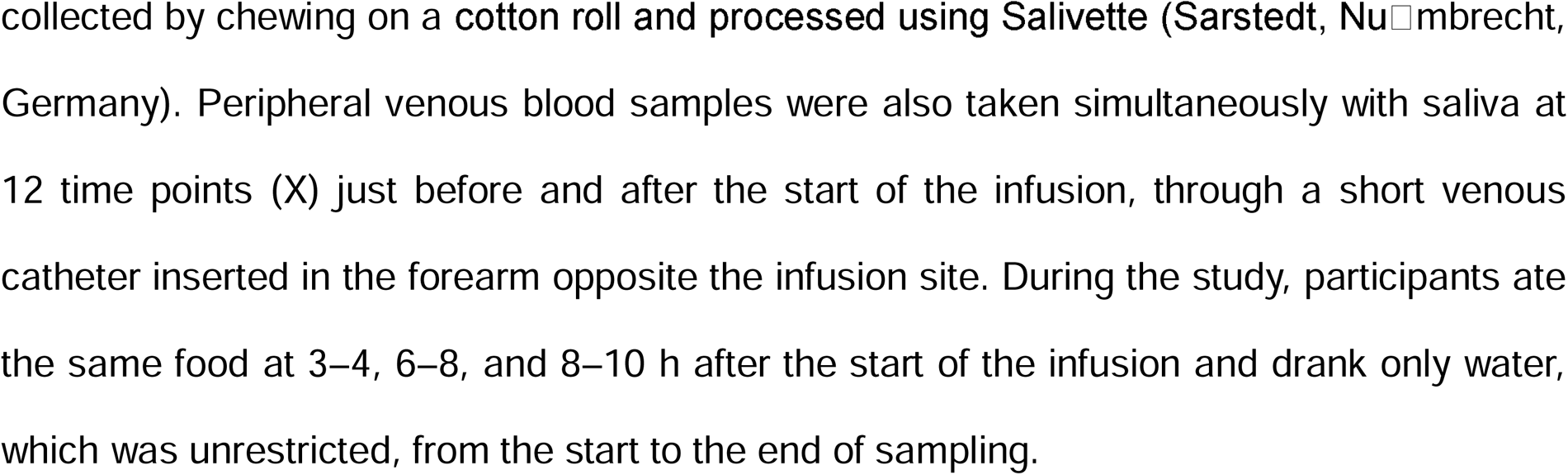
Time schedule of tedizolid study.

| Time (h) after<br>infusion | Infusion | Collection |  | Comment |
| --- | --- | --- | --- | --- |
|  |  | Saliva | Serum |  |
| 0 | Tedizolid phosphate<br>(Sivectro<sup>®</sup> 200mg) were<br>infused intravenously for<br>1 h. | X | X | Just before infusion |
| 0.25 |  |  |  |  |
| 0.5 |  | X | X |  |
| 0.75 |  | X |  |  |
| 1 |  | X | X | At the end of infusion |
| 1.25 |  | X |  |  |
| 1.5 |  | X | X |  |
| 2 |  | X | X |  |
| 2.5 |  | X | X |  |
| 3 |  | X | X |  |
| 3–4 |  | Food intake |  |  |
| 4 |  | X | X |  |
| 5 |  | X | X |  |
| 6 |  | X | X |  |
| 6–8 |  | Food intake |  |  |
| 8 |  | X | X |  |
| 8–10 |  | Food intake |  |  |
| 10 |  | X | X |  |

Saliva samples were taken at 15 time points (X) just before and after the start of the tedizolid infusion. To collect saliva samples, participants were asked to drink 50 mL of distilled water to promote saliva secretion before saliva collection. Subsequently, the saliva samples were
collected by chewing on a cotton roll and processed using Salivette (Sarstedt, Nußmberg, Germany). Peripheral venous blood samples were also taken simultaneously with saliva at 12 time points (X) just before and after the start of the infusion, through a short venous catheter inserted in the forearm opposite the infusion site. During the study, participants ate the same food at 3–4, 6–8, and 8–10 h after the start of the infusion and drank only water, which was unrestricted, from the start to the end of sampling.

## REFERENCES

1. Turner NA, Sharma-Kuinkel BK, Maskarinec SA, Eichenberger EM, Shah PP, Carugati M, Holland TL, Fowler VG. 2019. Methicillin-resistant Staphylococcus aureus: an overview of basic and clinical research. Nature Reviews Microbiology 17:203–218.

2. Kawasuji H, Nagaoka K, Tsuji Y, Kimoto K, Takegoshi Y, Kaneda M, Murai Y, Karaushi H, Mitsutake K, Yamamoto Y. 2023. Effectiveness and Safety of Linezolid Versus Vancomycin, Teicoplanin, or Daptomycin against Methicillin-Resistant Staphylococcus aureus Bacteremia: A Systematic Review and Meta-Analysis. Antibiotics (Basel) 12.

3. Cairns KA, Udy AA, Peel TN, Abbott IJ, Dooley MJ, Peleg AY. 2023. Therapeutics for Vancomycin-Resistant Enterococcal Bloodstream Infections. Clinical Microbiology Reviews 36:e00059–22.

4. World Health Organization (WHO) (2019) WHO consolidated guidelines on drug-resistant tuberculosis treatment. WHO G.

5. Agyeman AA, Ofori-Asenso R. 2016. Efficacy and safety profile of linezolid in the treatment of multidrug-resistant (MDR) and extensively drug-resistant (XDR) tuberculosis: a systematic review and meta-analysis. Ann Clin Microbiol Antimicrob 15:41.

6. Daley CL, Iaccarino JM, Lange C, Cambau E, Wallace RJ, Jr, Andrejak C, Böttger EC, Brozek J, Griffith DE, Guglielmetti L, Huitt GA, Knight SL, Leitman P, Marras TK, Olivier KN, Santin M, Stout JE, Tortoli E, van Ingen J, Wagner D, Winthrop KL. 2020. Treatment of Nontuberculous Mycobacterial Pulmonary Disease: An Official ATS/ERS/ESCMID/IDSA Clinical Practice Guideline. Clinical Infectious Diseases 71:e1–e36.

7. Garrabou G, Soriano À, Pinós T, Casanova-Mollà J, Pacheu-Grau D, Morén C, García-Arumí E, Morales M, Ruiz-Pesini E, Catalán-Garcia M, Milisenda JC, Lozano E, Andreu AL, Montoya J, Mensa J, Cardellach F. 2017. Influence of Mitochondrial Genetics on the Mitochondrial Toxicity of Linezolid in Blood Cells and Skin Nerve Fibers. Antimicrob Agents Chemother 61.

8. Wasserman S, Meintjes G, Maartens G. 2016. Linezolid in the treatment of drug-resistant tuberculosis: the challenge of its narrow therapeutic index. Expert Rev Anti Infect Ther 14:901–15.

9. Bandín-Vilar E, García-Quintanilla L, Castro-Balado A, Zarra-Ferro I, González-Barcia M, Campos-Toimil M, Mangas-Sanjuan V, Mondelo-García C, Fernández-Ferreiro A. 2023. Correction: A Review of Population Pharmacokinetic Analyses of Linezolid. Clinical Pharmacokinetics 62:1331–1331.

10. Mabilat C, Gros MF, Nicolau D, Mouton JW, Textoris J, Roberts JA, Cotta MO, van Belkum A, Caniaux I. 2020. Diagnostic and medical needs for therapeutic drug monitoring of antibiotics. European Journal of Clinical Microbiology & Infectious Diseases 39:791–797.

11. Pea F, Viale P, Cojutti P, Del Pin B, Zamparini E, Furlanut M. 2012. Therapeutic drug monitoring may improve safety outcomes of long-term treatment with linezolid in adult patients. J Antimicrob Chemother 67:2034–42.

12. Cojutti PG, Merelli M, Bassetti M, Pea F. 2019. Proactive therapeutic drug monitoring (TDM) may be helpful in managing long-term treatment with linezolid safely: findings from a monocentric, prospective, open-label, interventional study. J Antimicrob Chemother doi:10.1093/jac/dkz374.

13. Kawasuji H, Tsuji Y, Ogami C, Kimoto K, Ueno A, Miyajima Y, Kawago K, Sakamaki I, Yamamoto Y. 2021. Proposal of initial and maintenance dosing regimens with linezolid for renal impairment patients. BMC Pharmacology and Toxicology 22:13.

14. Kawasuji H, Tsuji Y, Ogami C, Kaneda M, Murai Y, Kimoto K, Ueno A, Miyajima Y, Fukui Y, Sakamaki I, Yamamoto Y. 2021. Initially Reduced Linezolid Dosing Regimen to Prevent Thrombocytopenia in Hemodialysis Patients. Antibiotics (Basel) 10.

15. van den Elsen SHJ, Oostenbrink LM, Heysell SK, Hira D, Touw DJ, Akkerman OW, Bolhuis MS, Alffenaar JC. 2018. Systematic Review of Salivary Versus Blood Concentrations of Antituberculosis Drugs and Their Potential for Salivary Therapeutic Drug Monitoring. Ther Drug Monit 40:17–37.

16. Davies Forsman L, Kim HY, Nguyen TA, Alffenaar J-WC. 2024. Salivary Therapeutic Drug Monitoring of Antimicrobial Therapy: Feasible or Futile? Clinical Pharmacokinetics 63:269–278.

17. van den Elsen SHJ, Akkerman OW, Jongedijk EM, Wessels M, Ghimire S, van der Werf TS, Touw DJ, Bolhuis MS, Alffenaar JC. 2020. Therapeutic drug monitoring using saliva as matrix: an opportunity for linezolid, but challenge for moxifloxacin. Eur Respir J 55.

18. Zailani NNB, Ho PC-L. 2023. Dried Blood Spots—A Platform for Therapeutic Drug Monitoring (TDM) and Drug/Disease Response Monitoring (DRM). European Journal of Drug Metabolism and Pharmacokinetics 48:467–494.

19. Seyfinejad B, Ozkan SA, Jouyban A. 2021. Recent advances in the determination of unbound concentration and plasma protein binding of drugs: Analytical methods. Talanta 225:122052.

20. Bolhuis MS, van Altena R, van Hateren K, de Lange WCM, Greijdanus B, Uges DRA, Kosterink JGW, van der Werf TS, Alffenaar JWC. 2013. Clinical Validation of the Analysis of Linezolid and Clarithromycin in Oral Fluid of Patients with Multidrug-Resistant Tuberculosis. Antimicrobial Agents and Chemotherapy 57:3676–3680.

21. Hara S, Uchiyama M, Yoshinari M, Matsumoto T, Jimi S, Togawa A, Takata T, Takamatsu Y. 2015. A simple high-performance liquid chromatography for the determination of linezolid in human plasma and saliva. Biomed Chromatogr 29:1428–31.

22. Anonymous. Pfizer 2005. Zyvoxid. Product information. Pfizer, New York, NY.

23. Moran GJ, Fang E, Corey GR, Das AF, De Anda C, Prokocimer P. 2014. Tedizolid for 6 days versus linezolid for 10 days for acute bacterial skin and skin-structure infections (ESTABLISH-2): a randomised, double-blind, phase 3, non-inferiority trial. Lancet Infect Dis 14:696–705.

24. Lee AS, de Lencastre H, Garau J, Kluytmans J, Malhotra-Kumar S, Peschel A, Harbarth S. 2018. Methicillin-resistant Staphylococcus aureus. Nature Reviews Disease Primers 4:18033.

25. Ruiz P, Causse M, Vaquero M, Casal M. 2019. *In Vitro* Activity of Tedizolid against *Mycobacterium tuberculosis*. Antimicrobial Agents and Chemotherapy 63:10.1128/aac.01939-18.

26. Brown-Elliott BA, Wallace RJ. 2017. *In Vitro* Susceptibility Testing of Tedizolid against Nontuberculous Mycobacteria. Journal of Clinical Microbiology 55:1747–1754.

27. Ruth MM, Koeken VACM, Pennings LJ, Svensson EM, Wertheim HFL, Hoefsloot W, van Ingen J. 2019. Is there a role for tedizolid in the treatment of non-tuberculous mycobacterial disease? Journal of Antimicrobial Chemotherapy 75:609–617.

28. Kumar K, Daley CL, Griffith DE, Loebinger MR. 2022. Management of *Mycobacterium avium* complex and *Mycobacterium abscessus* pulmonary disease: therapeutic advances and emerging treatments. European Respiratory Review 31:210212.

29. Iqbal K, Milioudi A, Wicha SG. 2022. Pharmacokinetics and Pharmacodynamics of Tedizolid. Clinical Pharmacokinetics 61:489–503.

30. Tsuji Y, Holford NHG, Kasai H, Ogami C, Heo YA, Higashi Y, Mizoguchi A, To H, Yamamoto Y. 2017. Population pharmacokinetics and pharmacodynamics of linezolid-induced thrombocytopenia in hospitalized patients. Br J Clin Pharmacol 83:1758–1772.

31. Inoue Y, Sato Y, Kashiwagi H, Nashimoto S, Sugawara M, Takekuma Y. 2023. Monitoring Salivary Concentrations of Tedizolid and Linezolid Using Rats. Eur J Drug Metab Pharmacokinet 48:387–395.

32. Tsuji Y, Numajiri M, Ogami C, Kurosaki F, Miyamoto A, Aoyama T, Kawasuji H, Nagaoka K, Matsumoto Y, To H, Yamamoto Y. 2021. Development of a simple method for measuring tedizolid concentration in human serum using HPLC with a fluorescent detector. Medicine (Baltimore) 100:e28127.

33. Elmongy H, Abdel-Rehim M. 2016. Saliva as an alternative specimen to plasma for drug bioanalysis: A review. TrAC Trends in Analytical Chemistry 83:70–79.

34. Aps JK, Martens LC. 2005. Review: The physiology of saliva and transfer of drugs into saliva. Forensic Sci Int 150:119–31.

35. Patsalos PN, Berry DJ. 2013. Therapeutic drug monitoring of antiepileptic drugs by use of saliva. Ther Drug Monit 35:4–29.

