## Supplementary figures and images for "High feasibility of salivary therapeutic drug monitoring in linezolid, but less in tedizolid: A single-dose study in healthy subjects"

### Figure S1

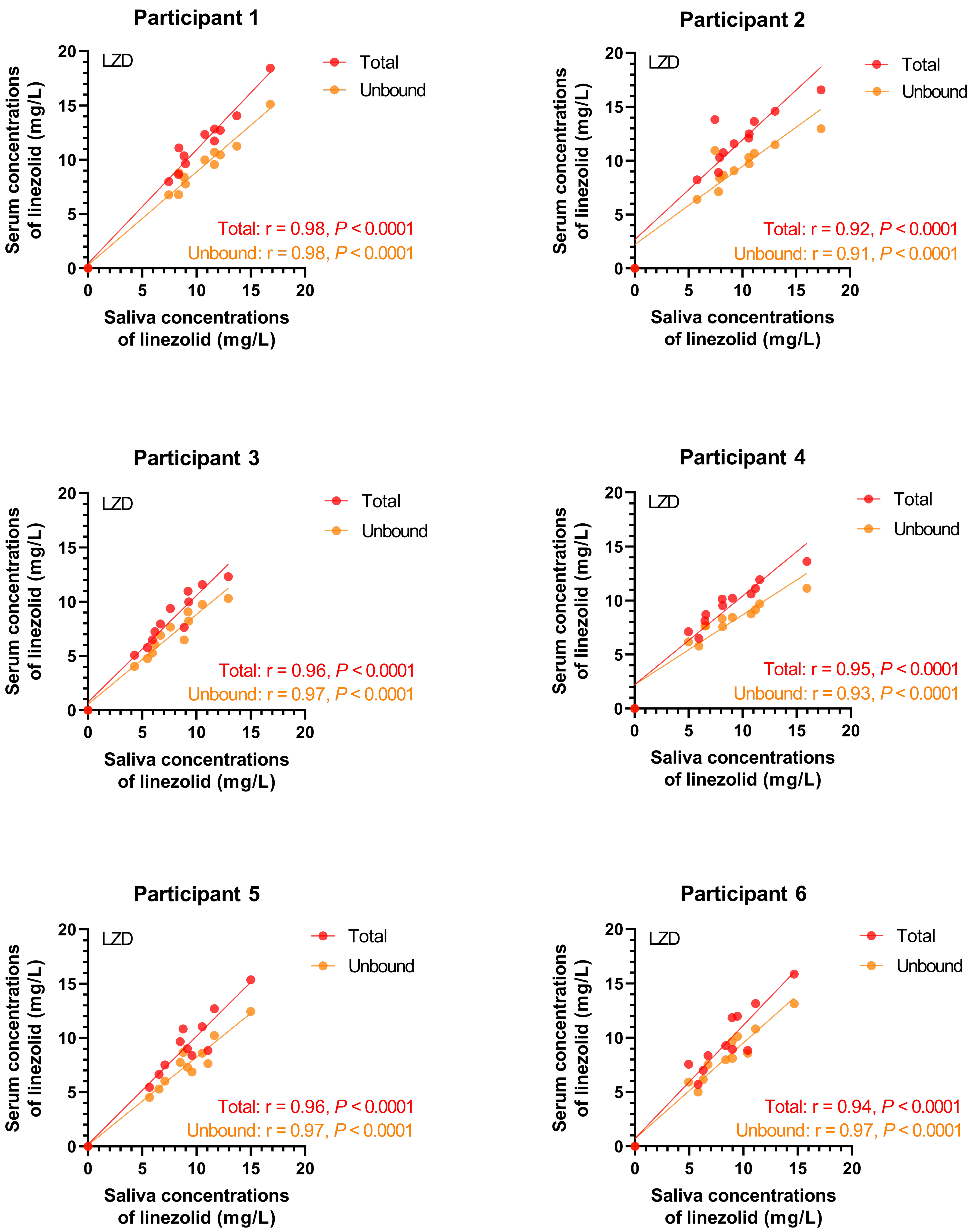

### Figure S2

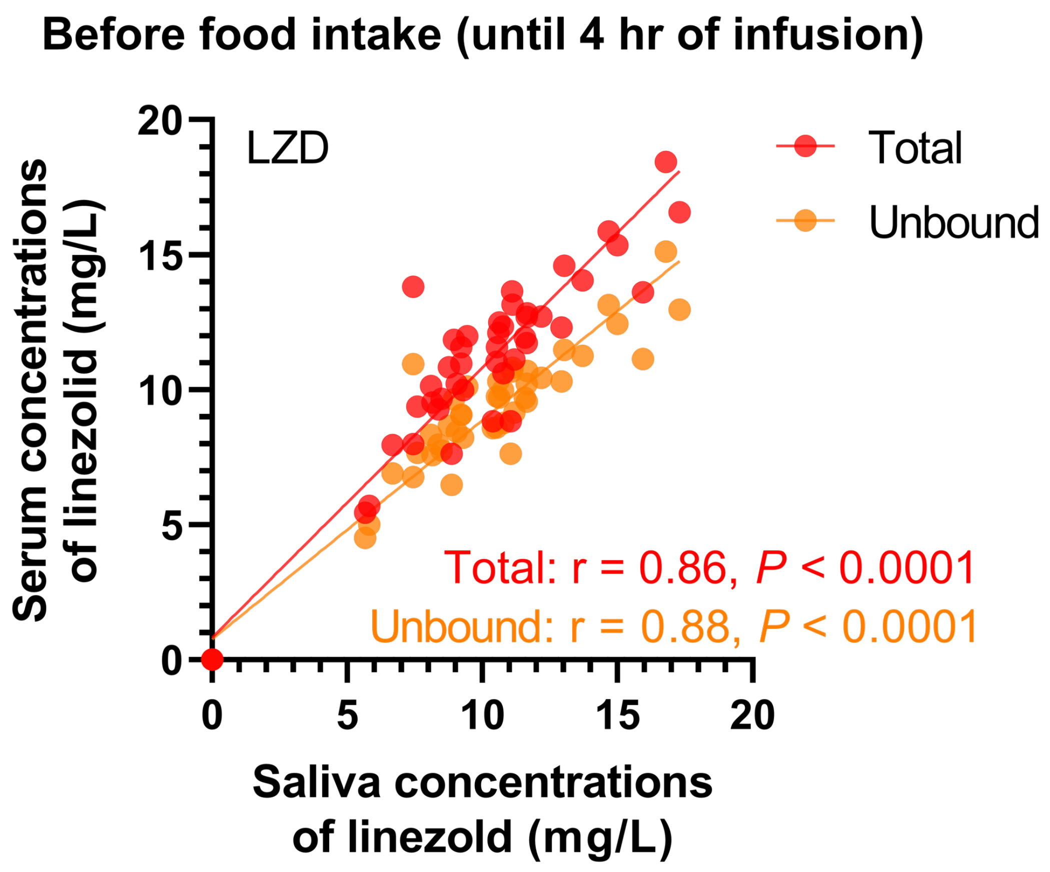

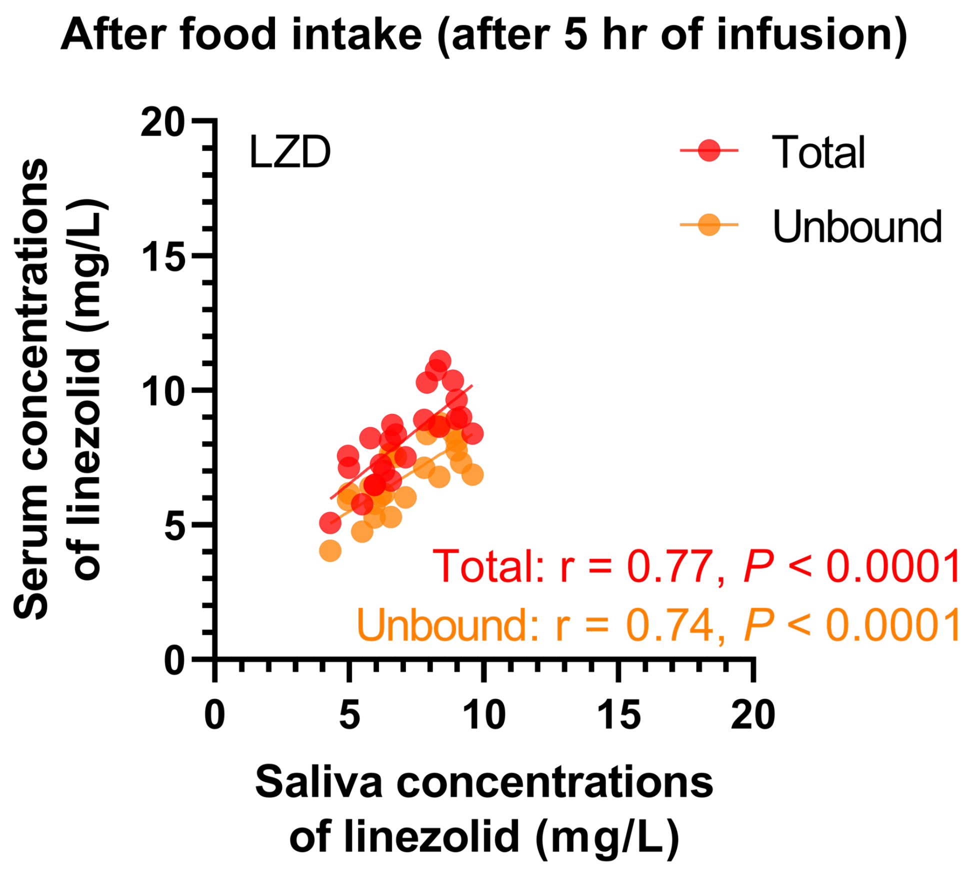

### Figure S3

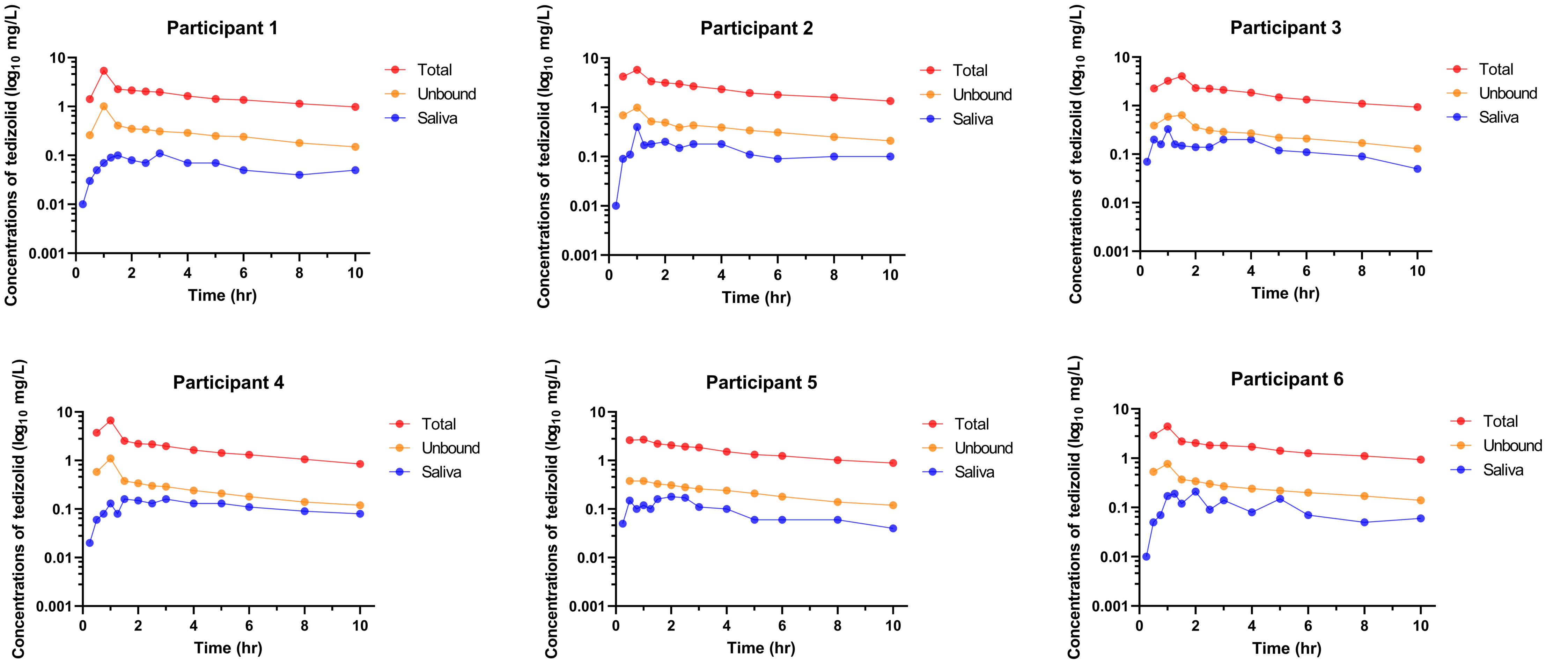

### Figure S4

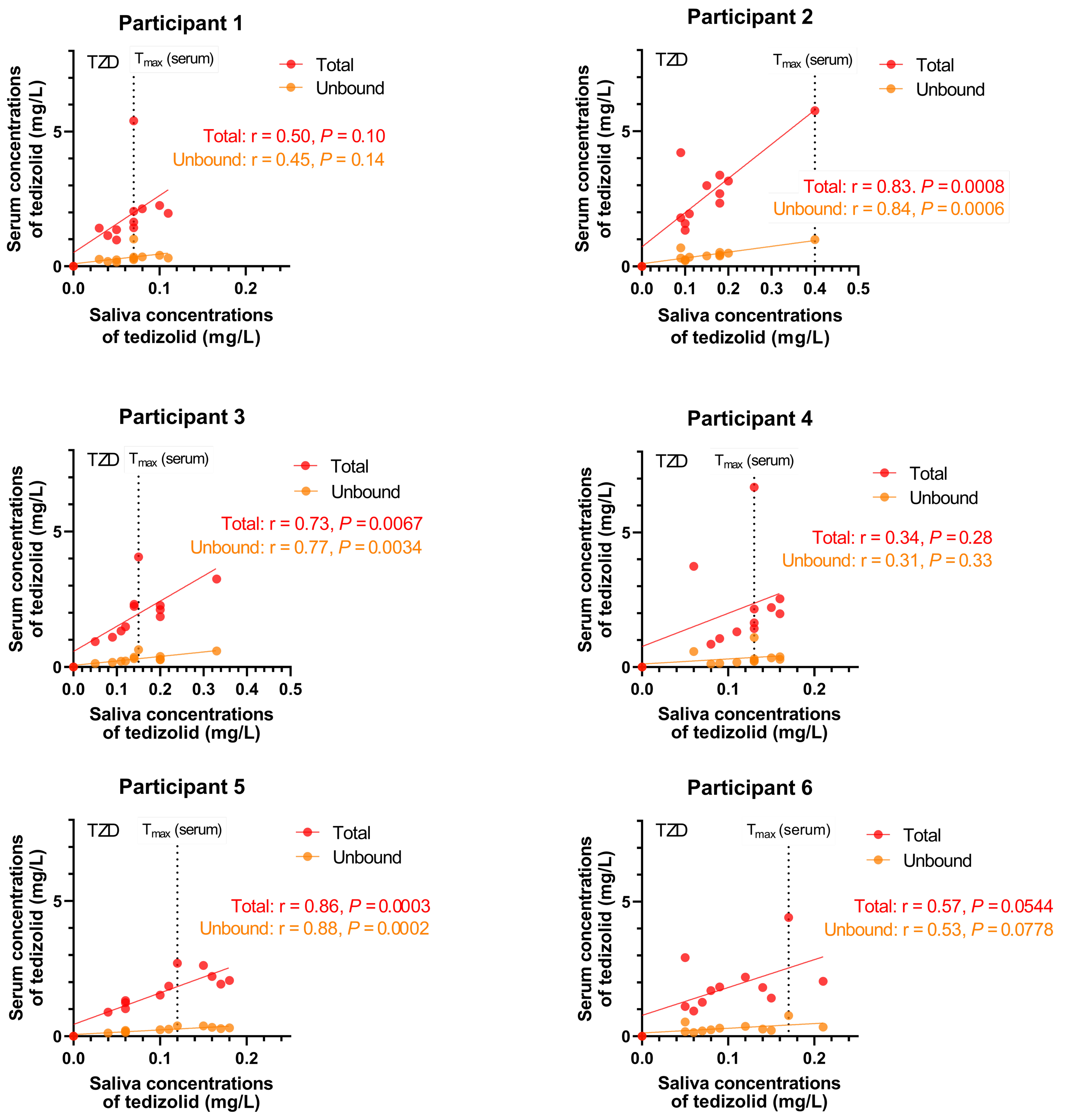

### Figure S5

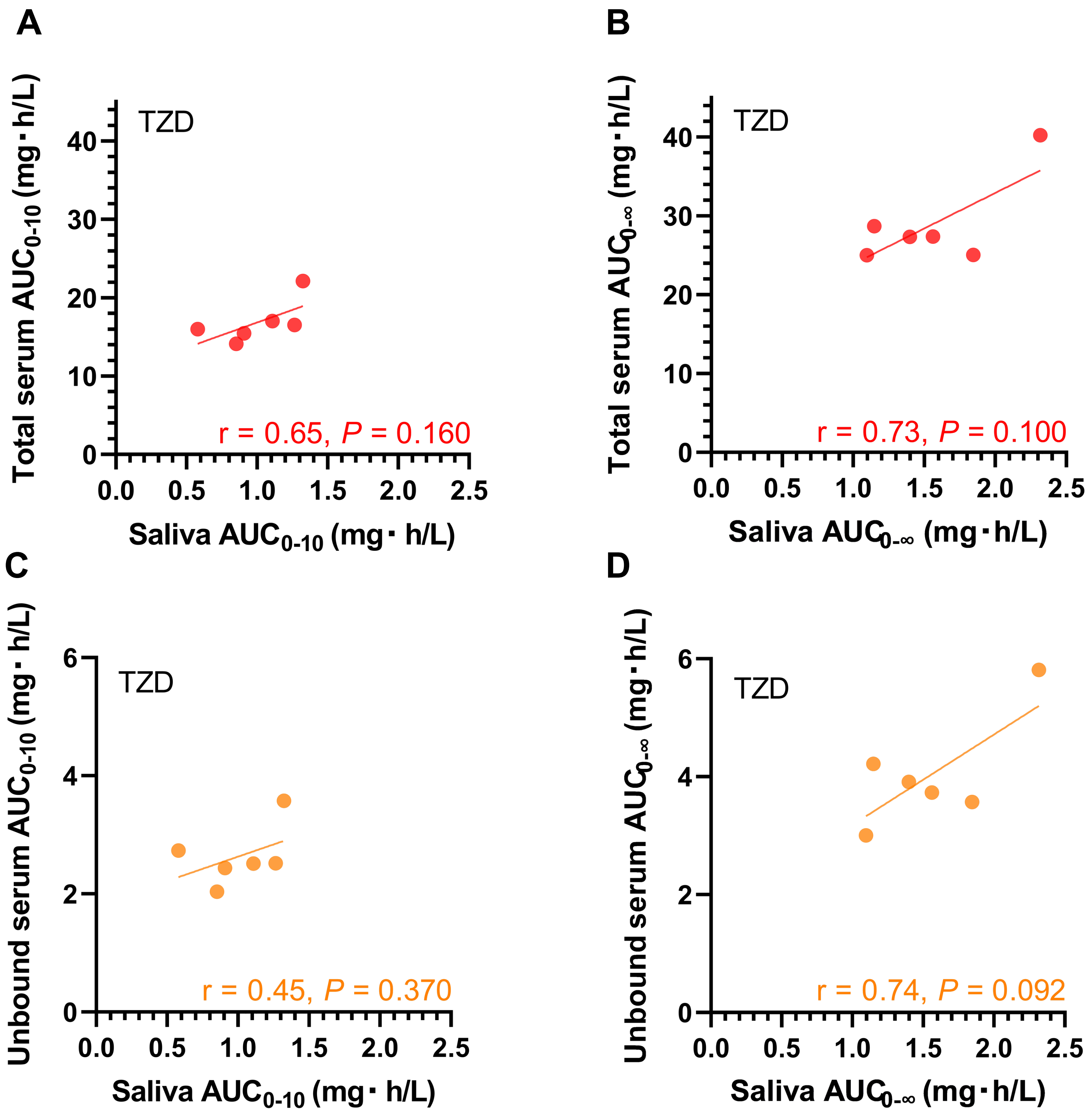
